# Evaluating the Perceived Utility of an Artificial Intelligence-Powered Clinical Decision Support System for Depression Treatment Using a Simulation Centre

**DOI:** 10.1101/2021.04.21.21255899

**Authors:** Myriam Tanguay-Sela, David Benrimoh, Christina Popescu, Tamara Perez, Colleen Rollins, Emily Snook, Eryn Lundrigan, Caitrin Armstrong, Kelly Perlman, Robert Fratila, Joseph Mehltretter, Sonia Israel, Monique Champagne, Jérôme Williams, Jade Simard, Sagar V. Parikh, Jordan F. Karp, Katherine Heller, Outi Linnaranta, Liliana Gomez Cardona, Gustavo Turecki, Howard Margolese

## Abstract

Aifred is a clinical decision support system (CDSS) that uses artificial intelligence to assist physicians in selecting treatments for major depressive disorder (MDD) by providing probabilities of remission for different treatment options based on patient characteristics. We evaluated the utility of the CDSS as perceived by physicians participating in simulated clinical interactions. Twenty psychiatry and family medicine staff and residents completed a study in which each physician had three 10-minute clinical interactions with standardized patients portraying mild, moderate, and severe episodes of MDD. During these scenarios, physicians were given access to the CDSS, which they could use in their treatment decisions. The perceived utility of the CDSS was assessed through self-report questionnaires, scenario observations, and interviews. 60% of physicians perceived the CDSS to be a useful tool in their treatment-selection process, with family physicians perceiving the greatest utility. Moreover, 50% of physicians would use the tool for all patients with depression, with an additional 35% noting they would reserve the tool for more severe or treatment-resistant patients. Furthermore, clinicians found the tool to be useful in discussing treatment options with patients. The efficacy of this CDSS and its potential to improve treatment outcomes must be further evaluated in clinical trials.

## 1. Introduction

Clinical decision support systems (CDSS) consolidate large quantities of clinical information to provide clinicians with actionable data to support medical decision-making and assist with managing treatment protocols (Sutton et al., 2020; Zikos and DeLellis, 2018). Increasingly, artificial intelligence (AI) algorithms are being integrated into CDSS, allowing for the deployment of predictive analytics by clinicians as part of routine practice (Sutton et al., 2020).

We created a CDSS which integrates an AI model that uses individual patient clinical and demographic characteristics to provide clinicians with remission probabilities for specific depression treatments (Benrimoh et al., 2020). We studied our CDSS at a simulation centre to evaluate its ease of use in routine practice, impact on the physician-patient interaction, and perceived utility. The two former points have been addressed in a previous publication (Benrimoh et al., 2020) and the latter is the focus of this paper. We were interested in how useful primary care physicians and psychiatrists would find the CDSS in assisting with their treatment decisions and in discussing these treatment options with patients.

The CDSS design was informed by important characteristics identified in discussions with physicians, including the simplicity of the interface and clinical utility of information displayed, as well as integration of the AI results into existing guidelines. It was important that the integration of the AI into the CDSS follow generalizable principles that, if validated, could be used to design similar tools. This included layering the AI predictions on top of existing best evidence guidelines (the CANMAT guidelines of the treatment of depression: Kennedy et al., 2016) and clearly labelling the AI predictions so that clinicians could consciously choose when to use them in their decision making. To improve interpretability (Benrimoh et al., 2018) and preserve physician autonomy, we designed our AI tool to provide reports detailing the key variables that informed each prediction and displayed the AI results as probabilities of remission, rather than suggestions or positive recommendations (see Mehltretter et al., 2020, for methodological details). The tool was also designed to facilitate shared decision making as a means of fostering patient autonomy and agency in the clinical encounter while positively supporting the clinician-patient relationship. Full details of the methodology followed in developing the CDSS, how it is differentiated from other CDSS, as well as the design of the simulation center study, can be found in Benrimoh et al. (2020).

Our main hypothesis was that clinicians would perceive the tool to be useful in shared decision-making with patients. We also hypothesized that primary care physicians (PCPs) and psychiatrists would use the tool differently due to their differing expertise and experience in the treatment of depression.

## 2. Methods

### 2.1. Participants and study design

The study sample consisted of intended users of the CDSS: primary care and psychiatry staff and residents. Twenty participating physicians were recruited for the study via social media and email. Participants provided informed consent and were compensated for their time. The study was approved by the Douglas Mental Health University Institute Research Ethics Board.

The study was conducted at the McGill Steinberg Centre for Simulation and Interactive Learning. The simulation centre provided nine standardized patients (SPs), professional actors trained to act as patients, who were compensated for their involvement. The use of SPs has been shown to be a valid reflection of patient experience (Beullens et al., 1997; Shirazi et al., 2011). SPs also ensure internal validity by allowing for each participant to respond to identical clinical scenarios. The simulation centre setup included a one-way mirror arrangement and an auditory monitoring system, allowing research assistants (RAs) to observe and listen to the simulated clinical scenarios.

Three 10-minute clinical scenarios - one each of a mild, moderate and severely depressed patient - were created by a clinician (D.B.), based on real patient data from the de-identified datasets on which the CDSS model was trained. Participants experienced the scenarios in a random order. Further details on the scenarios can be found in the supplementary methods and in Benrimoh et al. (2020).

Before the start of the simulation, we gave a short presentation to introduce participants to the basic principles of AI that our model uses. This was a structured presentation which can be found in the supplementary methods and included an explanation of the type of data used to train the model, the fact that a neural network was used, as well as the model metrics and the output of the model. Participants did not have significant prior knowledge of the AI or its development process. Participants then underwent a 10-minute training session with an RA which included teaching on navigating the tool. They were informed that the ‘patients’ had used the tool to complete questionnaires prior to their session, but that they had a limited understanding of how the AI model operated.

Participants conducted the 10-minute clinical consultations with the SP as per their usual practice and integrated the CDSS as they saw fit. It was suggested, but not mandated, that they spend five minutes interviewing the patient followed by five minutes using the CDSS. Within the CDSS, participants had access to the patient’s questionnaire results and the treatment algorithm with the integrated predictive model. The provided laptop was positioned at a 45 degree angle towards the participant in order to make the screen visible to researchers observing the interaction, though participants were free to move the laptop. Participants were informed that the clinical scenarios were ten minutes long, and had access to a clock. Following each clinical scenario, participants completed a questionnaire regarding their use of the CDSS model. After the three scenarios, physician participants were interviewed by RAs using a standard semi-structured interview with predefined questions including both open-ended and closed, specific questions (see supplementary methods) and completed a questionnaire about their experience using the model, as well as a short quiz to assess their familiarity with the CANMAT 2016 guidelines for depression treatment. Standardized patients were interviewed in an unstructured manner as a group at the end of each testing day and their observations were recorded in writing by RAs. This was done in order to obtain their immediate impressions about their interaction with clinicians during the testing day. In addition, given that SPs sometimes changed between days, this ensured that all SPs were able to provide feedback.

In order to improve reporting quality, we endeavored to report our results in line with the suggested amendments to the STROBE guidelines for the reporting of simulation-based research (Cheng et al., 2016). Custom questionnaires were used due to the novelty of the CDSS. Participants were surveyed about their previous simulation experience.

### 2.2. Quantitative analysis

The descriptive results from participant self-report questionnaires were generated using R v.3.3.2 and visualized using the ggplot2 v.3.2.1 package.

### 2.3. Qualitative analysis

The qualitative data consisted of the written and interview feedback from the participants and standardized patients and the RAs’ written observations on the clinical scenarios. Interviews were not recorded but RAs kept extensive notes and then transcribed these notes into digital spreadsheets. Coding was done using digital spreadsheets. Initial themes were brainstormed prior to data analysis and interpretation as a means of unbiasedly commencing the codification of the data; nine themes emerged from this initial effort: 1) interpretability of the AI report; 2) degree of trust in AI; 3) user experience for the patient; 4) user experience for the clinician; 5) the tool’s impact on the physician-patient interaction; 6) the potential role and use of the AI tool in practice; 7) suggested tool improvements; 8) user interface; and 9) situations in which the clinician felt they would use the tool. Each theme had a number of related sub-themes. Using an inductive thematic analysis approach allowed the data to be coded without trying to fit it into a pre-existing coding frame or the researcher’s analytic preconceptions, and allowed the themes identified to be strongly linked to the data themselves (Braun and Clarke, 2006).

The investigator triangulation method was employed during the qualitative analysis, in which multiple investigators compared individually coded qualitative data to reduce bias (Archibald, 2016). RAs independently read and coded excerpts of the data into subheadings of the thematic table. The source of the excerpt and the scenario from which it was extracted was noted. Four RAs were each assigned the data corresponding to ten participants, such that each participant’s data was independently coded by two RAs. An additional independent coder compiled all the excerpts. This data was condensed and redundancies eliminated by collapsing some of the themes and rearranging the subheadings. The RAs then independently reread and coded all of the data into a final summary table. This stage ensured that any data that had been missed in earlier coding stages could be added, and also validated the new themes in relation to the full data set. Triangulation of qualitative data sources involved the comparison of observational data with interview data, allowing the analysis of different perspectives. This approach offered a greater insight into the relationship between inquiry approach, data sources and the phenomenon under study.

## 3. Results

### 3.1. Quantitative analysis

#### 3.1.1. Sample description

Our sample comprised 11 psychiatrists (8 staff, 3 residents) and 9 PCPs (6 staff, 3 residents). Mean age of participants was 39.5 years (SD 13.3). Most participants were trained in Quebec (*n* = 17), and ranged from residents to staff with decades of experience (see Table 1). Clinical practice environments included both inpatient and outpatient services, and there was a wide range in the self-estimated number of patients treated for MDD per month (Table 1). Responses to questions related to current clinical practices, the potential role for clinical decision aids and AI technologies, as well as which treatments were prescribed during the simulation sessions can be found in Supplementary Table 1.

**Table 1.** Demographics (*n* = 20)

| Demographic | Response Options | Frequency |
| --- | --- | --- |
| Gender | Female | 13 (65%) |
|  | Male | 7 (35%) |
| Specialty | Primary Care Physician (PCP) | 9 (45%) |
|  | Psychiatrist | 11 (55%) |
| Training<br>*Where were you trained?* | Quebec | 17 (85%) |
|  | Rest of Canada | 3 (15%) |
| Resident Level<br>*If you are a resident, what is your level?* | N/A (staff clinicians) = 14 |  |
|  | PGY1 | 1 (5%) |
|  | PGY2 | 2 (10%) |
|  | PGY3 | 3 (15%) |
| **Staff Years Experience**<br>*If you are a staff member, how many years of experience do you have?* | **Residents** | **6 (30%)** |
|  | **0-5** | **4 (20%)** |
|  | **6-10** | **2 (10%)** |
|  | **11-15** | **2 (10%)** |
|  | **16-20** | **4 (20%)** |
|  | **21+** | **2 (10%)** |
| **Environments of Practice**<br>*Which environment(s) do you practice in? (Check all that apply.)* | **Hospital Outpatient Department** | **10 (50%)** |
|  | **Inpatient Service** | **8 (40%)** |
|  | **Outpatient Clinic** | **12 (60%)** |
|  | **ER** | **4 (20%)** |
|  | **Consult Liaison** | **1 (5%)** |
|  | **Specialized Mood Disorder Service** | **1 (5%)** |

### 3.2. Descriptive results

Question responses were recorded using Likert scales with the answer options “strongly disagree”, “somewhat disagree”, “unsure”, “somewhat agree”, and “strongly agree”. Here, “disagreed to a degree” and “agreed to a degree” or “to some degree” indicate that the “somewhat” and “strongly” results have been combined.

60% of participants agreed to some degree that the model was useful in making treatment decisions. More PCPs appeared to feel that the model was useful in making treatment decisions compared to psychiatrists (Figure 1).

**Figure 1.**
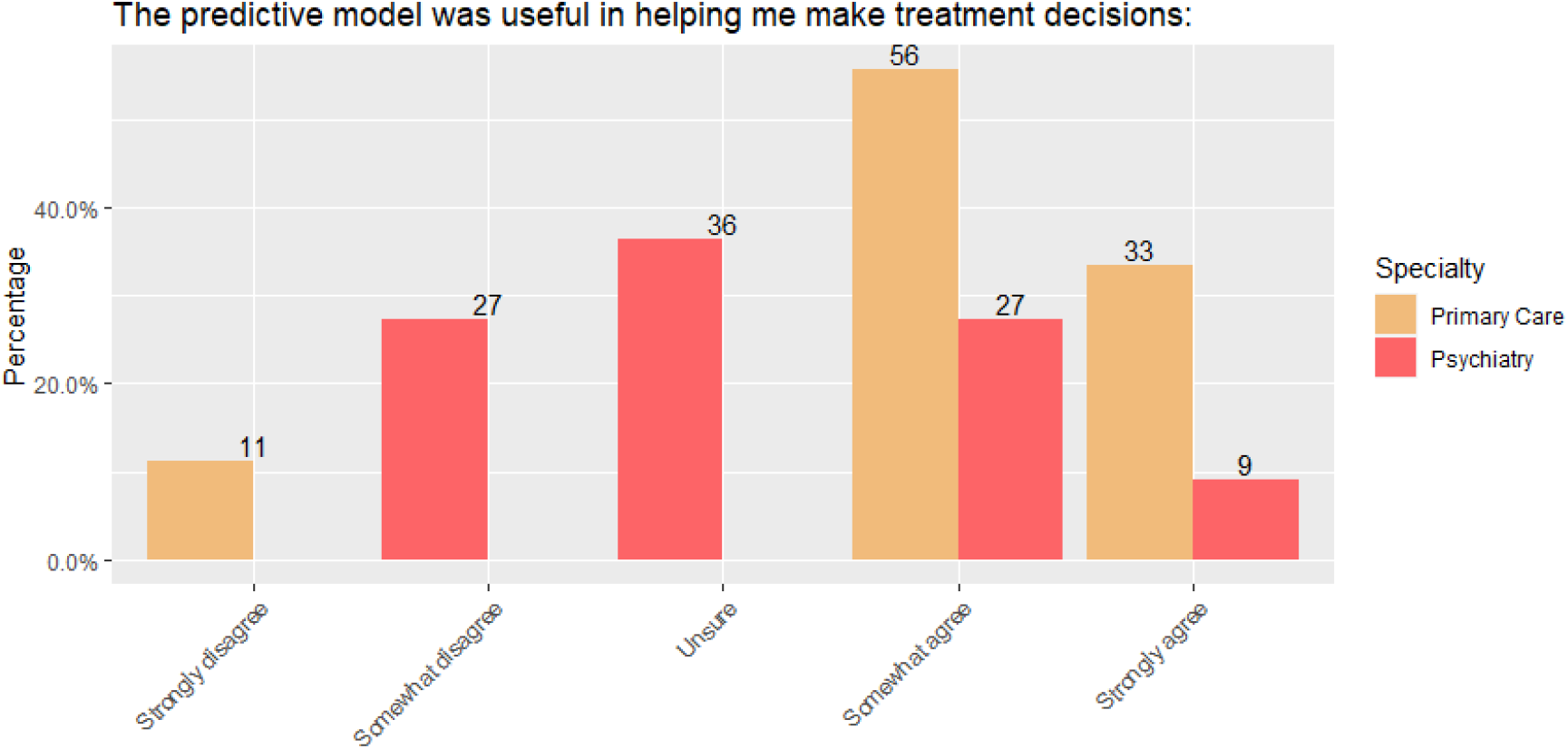
Ratings of the usefulness of the model in making treatment decisions.

Tests of statistical significance are not included for these figures because of the small sample size which means the study is underpowered compared to the number of comparisons shown. These figures are meant to help assess possible trends which should be replicated in larger samples in future studies.

70% of participants described the probabilities produced by the model as “reasonable”, while 15% described them as “too pessimistic” and the remaining 15% as “too optimistic” (Figure 2).

**Figure 2.**
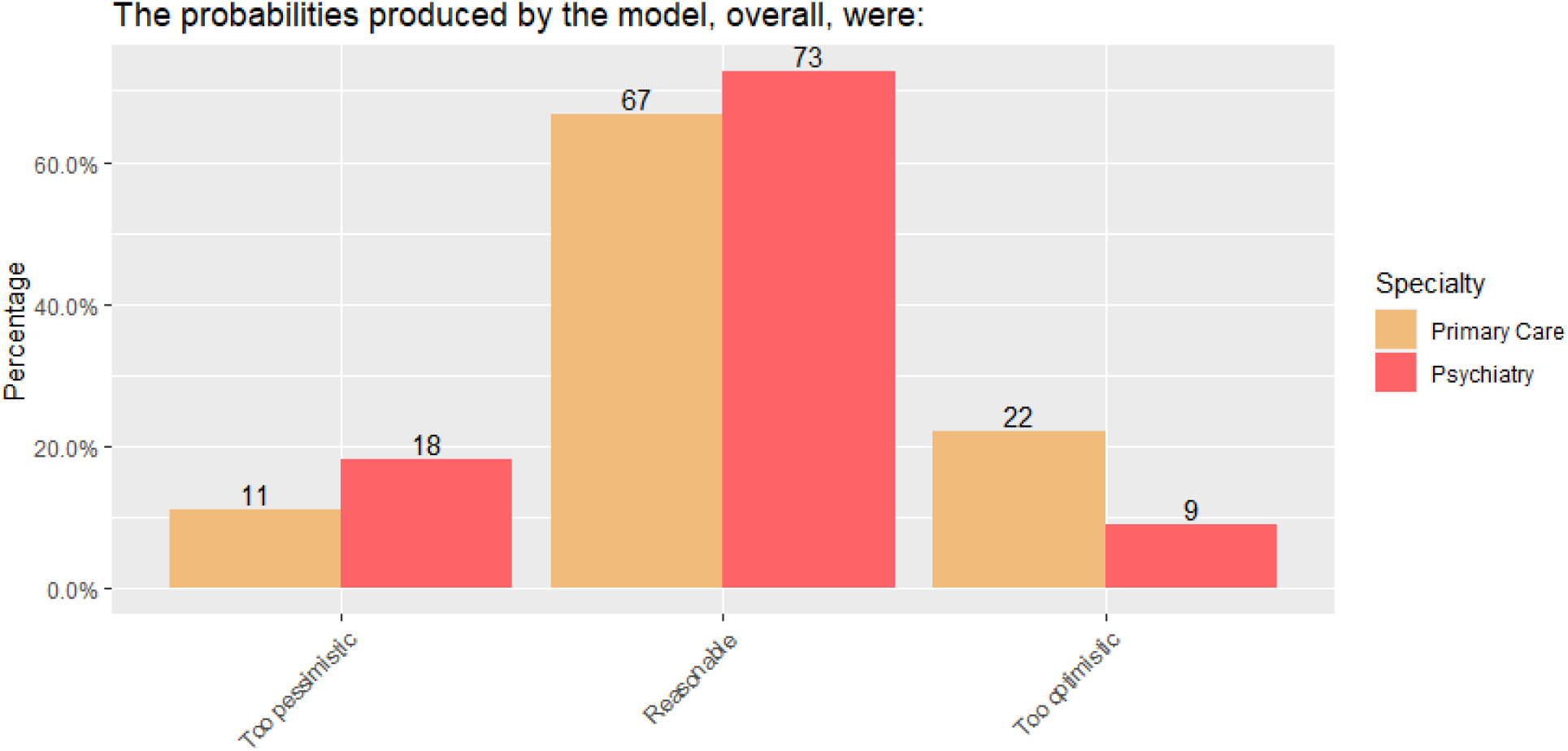
Ratings of perceived reasonableness of the model.

Participants were asked to rate their trust in the model on a scale of 1-5 based on each clinical experience. From the first session to the third session, the proportion of higher ratings of trust increased (Figure 3).

**Figure 3.**
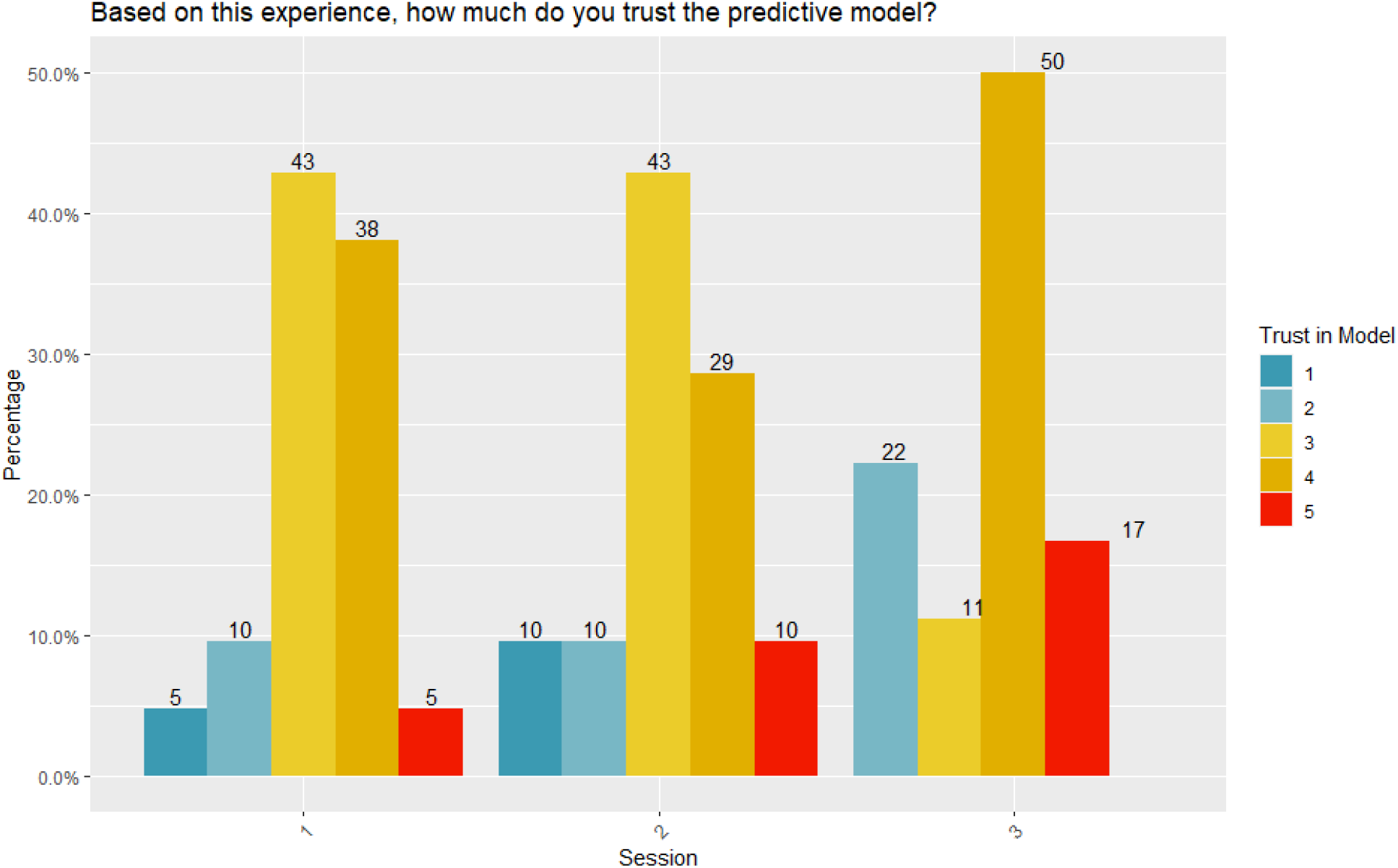
Ratings of trust in the model across simulation sessions (scale of 1 to 5, 1 being “very little” and 5 being “very much”)

Participants were also asked how much they felt the predictive model helped them make a treatment decision on a scale of 1-5. Across sessions, it appears that there is a shift towards increasing proportions of higher ratings on the scale (i.e., greater feeling that the model helped make the treatment decision) from session one to three (Figure 4).

**Figure 4.**
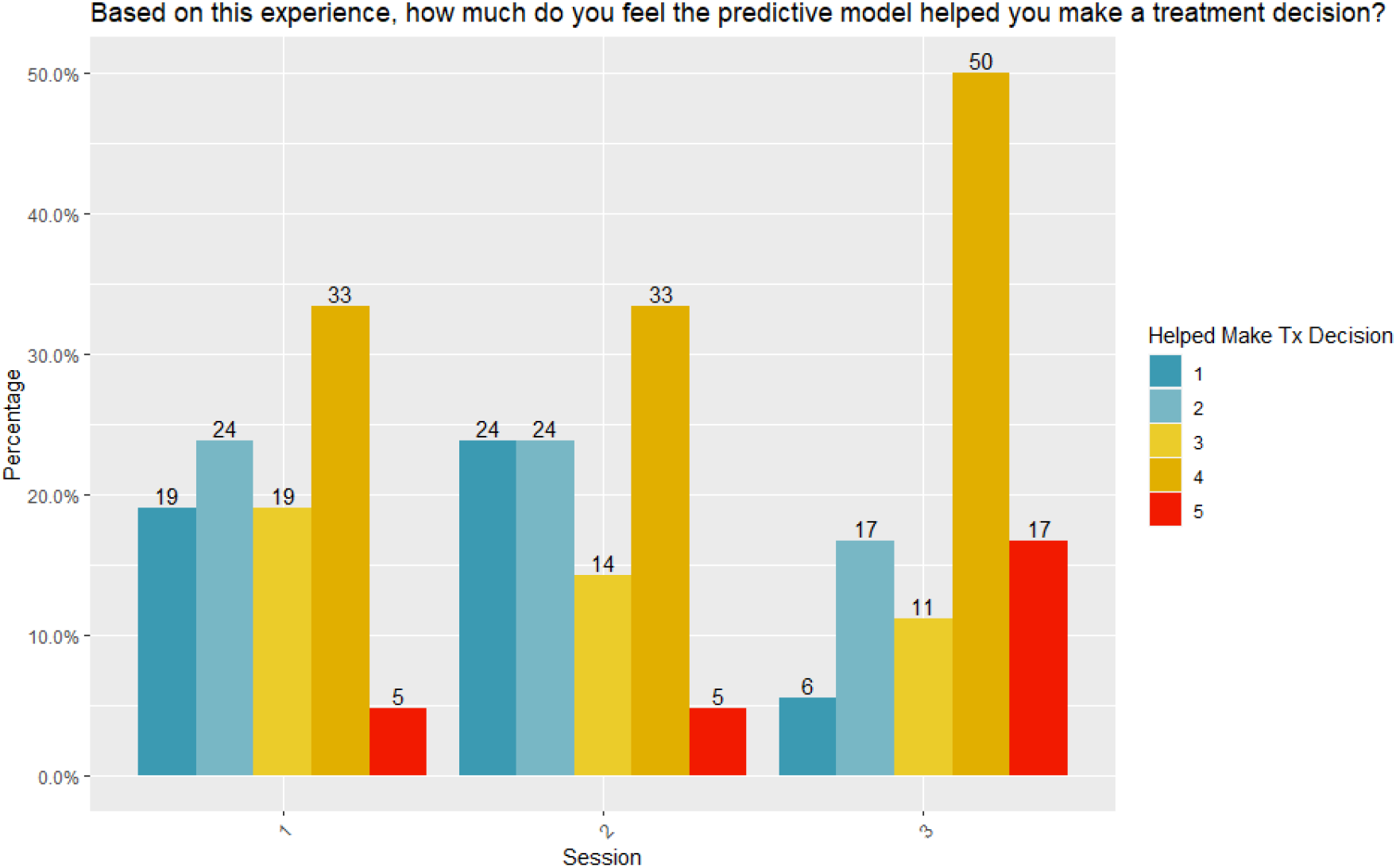
Ratings of the model’s helpfulness in making treatment decisions across simulation sessions (scale of 1 to 5, 1 being “very little” and 5 being “very much”)

In addition, nearly half (48%) of the time, physicians chose one of the top two treatments predicted by the AI (see supplementary materials for analysis and discussion of predictors and correlates of physician selection of treatments consistent with AI-defined optimal treatments). Results of the CANMAT quiz can also be found in the Supplementary Materials.

### 3.4. Perceived Utility - Qualitative Analysis

Our qualitative analysis provided additional insight into how physicians felt using our tool for the first time and their perceptions of its utility. After reading through the qualitative data, four RAs reduced the original nine themes to the following four in order to reduce the overlap between excerpts: interpretability of the tool, impact on treatment decision and clinical practice, trust and understanding of AI, and impact on physician-patient interaction. Each of the four themes had a number of sub-themes (Supplementary Table 3 and Supplementary Figure 2). Data presented will focus on the subset pertaining to the perceived utility of the tool. As a general note, RAs observed that use of the tool became progressively more integrated and that it was utilized more confidently across trials. Physicians reported that the tool was easy to use, but many nonetheless felt they would have benefited from more practice. Moreover, a shared sentiment among several physicians was that in their usual clinical practice, they would likely review some of the information presented in the tool prior to a session with their patient in order to have more time to digest the information.

#### 3.4.1. Trust, Understanding, Feelings about AI

Participant trust and belief in the AI predictive model was generally positive. Of the four participants who explicitly mentioned “trust” or “faith” with respect to the AI, two reported that they trusted the tool, one wondered about how much to trust the remission probabilities, and one reported having less trust in the AI after seeing one of the important features behind a prediction (which they did not consider to be predictive) but felt that the use of this tool would be especially beneficial in treating younger patients and gaining their credibility. Another participant reported that the use of AI in psychiatry could be beneficial by introducing some objectivity. Conversely, one participant worried about the integration of AI models in healthcare, “AI interprets data, but people are not data”.

Furthermore, six participants requested more evidence behind the AI model, specifically the datasets informing the remission probabilities. Of those, two expressed that if more information about the algorithmic reasoning behind treatment decisions had been available, they would have trusted the model more.

Four participants expressed they had a limited understanding of how the AI predictive model functioned, but were keen to integrate it into their treatment decisions. The comfort level of the participant with the AI model, reported via RA observations, was aligned with how well they were able to explain the model to the SP. The terminology used to describe the tool to the SPs varied greatly, including “tool”, “new technology”, “the algorithm”, and “the computer”. Interestingly, only 25% of participants used the term “AI” to describe the model, with another 25% preferring to describe it, for example, as a tool that “makes decisions based on data from many other individuals”.

#### 3.4.2. Impact on Treatment Decision and Clinical Practice

Participants discussed the impact of the model on their treatment decision. Roughly half (45%) of the responses were distinctly positive, describing the potential of the tool to transform practice or diversify treatment approaches. One participant noted that the CDSS “helps you to either choose a specific antidepressant medication or at least narrow down your range of choices to a few ideal candidates.” 25% of participants reported no impact on treatment decisions from the tool due to greater confidence in their own clinical judgment, perceived minimal differences in projected outcomes between suggested treatments, and/or noting that treatments suggested by the model were already in line with their prior treatment plan. Negative comments centered around the interference of the tool in the physician-patient relationship, or, for one participant, on the perception that the tool focused too greatly on medication.

A recurring theme was related to the significance of remission rate percentages offered by the tool: if the probabilities were felt to be clinically significant, participants described a greater impact on their treatment decision: “normally [I] wouldn’t prescribe a medication for [this] case [… but I will] because the percentage is quite high so I do think that it is worth trying the medication”; this is juxtaposed with the perception of little to no impact on treatment decision if the differences between percentages was interpreted as insignificant “[a] small difference in percentages is not going to change how I practice”. Participants described their perceived potential value of the tool in several ways: as a potential way to save time (10%); to confirm/suggest treatment options (15%); as a centralized tool for guidelines (10%); as a source of extra information about treatments (15%); as useful for displaying symptoms over time (5%, though it should be noted that a longitudinal data element was not included in this study); and as a way to explain to patients why a particular treatment is being chosen (15%). The value of gaining more familiarity with the model was also apparent from participant comments: “since this was the first time using it I did tend to stick to what I would usually prescribe, however I can see that if I got used to using it regularly as part of my residency training that I would probably use different treatment options”.

#### 3.4.3. Communicability and Interpretability of the CDSS’s Results

The key to the clinical utility of a tool aimed at supporting shared decision making is the ability of physicians to communicate results and their impact on decision-making to patients, and, relatedly, physician understanding of the tool. 40% of physicians made reference to the benefit of being able to communicate the motivation for a treatment decision to the patient, “especially to give patients concrete numbers about their remission probabilities”. As one physician observed, “One good thing was you could explain to the patient why you are choosing the treatments that you are choosing.” 20% of participants expressed wanting more information about the source of the remission probabilities, either about the model itself, the clinical data that trained the deep learning model, or about how the variables considered by the model impacted the remission prediction for an individual patient.

## 4. Discussion

In order to support clinical decision making, a CDSS must provide high-quality, clinically useful information that is relevant to the individual patient, and is accessible, interpretable, operational and actionable for the clinician (Sim et al., 2001). Additionally, the effort expended by the physician to use the system must not be perceived as excessive (Wendt et al., 2000). In this study of an AI-powered CDSS for depression treatment with a sample of intended users - primary care and psychiatry staff and residents - we aimed to evaluate perceived utility of the tool and perceived impact of the tool on clinical decision-making, and potential differences in perceived utility between PCPs and psychiatrists. In the supplementary material, we also discuss drivers of physician prescription of treatments consistent with those predicted by the AI model as having the highest likelihood of success. Conducting this study using simulated patients created a safe and controlled environment to measure the tool’s impact on the treatment decision-making process (Benrimoh et al., 2020).

In a previous paper reporting on this dataset (Benrimoh et al., 2020) we demonstrated that physicians felt the tool was feasible to use in a clinical interaction and did not have significantly deleterious effects on the patient-clinician interaction. Once a CDSS has met the basic requirement of being easy to use within a reasonable time frame (in our study, in roughly five minutes within a ten-minute interview), can be conceivably worked into the clinical workflow, and seems to garner the trust of most users, the next question to be answered is: do physicians find the tool to be useful with respect to its intended goal of assisting in treatment decisions? In our study, physicians were free to use or ignore CDSS predictions and to choose treatments they saw fit, providing us with an opportunity to use a mixed-methods approach to investigate their perceived utility of the system in a controlled environment where their interactions with the system and with patients could be observed, but where their actions within those interactions were not directed (i.e. they were not forced to use or pay attention to all or part of the tool).

Overall, the majority of participants found that the model was trustworthy and useful in making treatment decisions. Both trust and perceived usefulness in making treatment decisions increased over time, indicating, in line with the qualitative results, that improved familiarity with the CDSS and the resulting increased comfort allowed clinicians to better understand and integrate it into their approach, improving its perceived utility. Feedback from the participating physicians revealed differences between PCPs and psychiatrists. PCPs have been found to perceive the treatment of patients with depression as challenging, feeling that these patients place a high demand on their psychological resources (McPherson and Armstrong, 2012), with higher ratings for utility by PCPs. This sense that depressed patients present a treatment challenge, combined with the fact that PCPs may not be as familiar with the guidelines or the available range of medications as specialists, may have driven our finding that PCPs are more likely to find the model useful in making treatment decisions.

The mixed-methods approach of this study yielded qualitative data that can further nuance our understanding of perceived utility. The impact of the CDSS on treatment decision-making was seen as positive by a significant proportion of participants. Some participants noted the availability of questionnaire data and the predicted remission probabilities as being useful in shared decision making with patients, with some physicians specifically noting that they would use the remission probabilities to help patients understand possible treatment choices. Other elements of potential utility were noted, with some responses suggesting value from potential time savings as well as the ability to centralize information about treatments which can assist in the review of different options available. The ability to use the CDSS to “narrow down” the list of possible treatments was also highlighted. Taken together with the willingness to consider CDSS predictions demonstrated by participants (for example, in the quote described above where a participant physician was willing to consider prescribing a medication when they normally would not), and the fact that treatments chosen agreed with the model 48% of the time, this suggests that clinicians see the tool as being a useful aid to decision, but not as a replacement to their clinical judgement, especially when the predicted differences between treatments were small. Indeed, a number of participants chose to favor their own judgment over that of the CDSS, citing their greater confidence in their own experience. These results are in line with the design philosophy behind the CDSS, in that the tool is envisioned as an aid to clinicians and patients in shared decision making and in facilitating the use of measurement-based, algorithm-guided care, but not as a tool meant to ‘hijack’ clinical decision making.

One key result apparent in the qualitative data which was reflected in the quantitative results is the feeling on the part of physicians that greater time to work with and be exposed to the application, as well as an improved understanding of the AI, would improve the ease of use and comfort with the tool, increase trust in the tool, and would also likely increase its perceived utility for assisting clinical decision-making. For example, as discussed above, one participant physician mentioned that they chose to select the treatment they would usually prescribe during the session, but felt that with continued use of the tool they could see it helping them expand the treatment options they would consider. This result coheres with the finding in the quantitative data that the perceived utility of the model increased over the course of the three sessions. This information is critical because it will influence the design of training materials and procedures, ensuring that longer training periods as well as more comprehensive training materials are provided. In addition, in future clinical studies, the prediction that greater familiarity with the tool will yield increased trust in or utilization of CDSS treatment predictions could be tested.

## Limitations

Our findings should be interpreted considering several limitations. The small sample size limits the generalizability of the results and may bias the quantitative analysis (Moineddin et al., 2007), and also prevented the use of extensive testing for significance because of lack of power. As such, trends observed here should act as results to be replicated in future, larger clinical samples. In addition, the lack of a control group makes it impossible to know how physicians would have acted without access to the CDSS. However, the main purpose of the current study was to establish the ease of use and perceived utility of our CDSS. Future work in clinical populations will assess the feasibility, safety, clinical utility, and effectiveness of the CDSS and seek to replicate the current results in larger clinical samples. Given the limitations on time in the simulation center, participants were provided with only brief training without opportunity for repeated practice sessions. We observed that participant scores that rated dimensions of trust, ease, and helpfulness improved on average from the first trial to the last trial, suggesting that, as expected, comfort level with the CDSS increases as a function of practice. Moreover, participants expressed an interest in more extensive training and practice sessions, as well as more extensive information about the AI model. These will be provided in future clinical studies alongside extended training, but the limited training in this study may have impacted perceived utility. Finally, a simulation center is not a real clinical setting and testing in real clinical environments will be required to replicate and verify clinician perceived utility and verify validity of these results in usual clinical environments. A central question remains whether clinicians will use and continue to use this tool in real clinical practice, or if use will experience unexpected barriers or drop off with time; these questions are beyond what can be answered in a simulation setting and require further clinical research.

## Conclusion

We present preliminary findings of perceived clinical utility of a novel AI-enabled CDSS for physicians treating patients with depression. Overall, physicians found the system useful and beneficial during shared decision-making with patients, with PCPs having the greatest perceived utility. Physicians perceived the tool to be more useful with repeated use, consistent with their comments that greater training and time with the tool would likely increase their perceived utility. This CDSS presents a new opportunity to use readily available patient data to personalize treatment choice at the point of care, and preliminary results indicate that physicians are able and willing to use this kind of tool to support their decision making. Further advances in AI interpretability as well as improved training regimens for physicians should help improve trust and in turn use of AI results, and establishment of this kind of tool in the treatment of depression may lead to applications in other areas of mental health. The use of a mixed methods approach as well as the simulation center was useful in providing information that could benefit further development and improve training for participants in future studies. Clinical trials are furthermore required to assess the effectiveness of this tool in improving mental health outcomes. These trials will help determine the utility of the CDSS from the patient’s perspective, which is necessary to build a tool that delivers care aligned with patients’ needs and preferences. Creating a CDSS via a patient-centric approach has the potential to improve the support provided to patients and empower them to participate in their own care.

## Data Availability

Data is not publicly available. Interested parties should contact the authors.

## Acknowledgements

We would like to acknowledge the Steinberg Centre for Simulation and Interactive Learning for the helpfulness of their staff in assisting with the execution of this study, as well as the standardised patients who participated in the study, for their excellence and the quality of their feedback.

