## Supplementary Material for "Evaluating the Perceived Utility of an Artificial Intelligence-Powered Clinical Decision Support System for Depression Treatment Using a Simulation Centre"

Supplementary Materials

Supplementary Methods

**Participants and Study Design**

The study sample consisted of the intended users of the CDSS: primary care and psychiatry staff and residents. A total of twenty participants were recruited for the study via social media, email and announcements three months prior to the start of the study. All participants provided informed consent and were compensated for their time. The study was approved by the Douglas Mental Health University Institute Research Ethics Board.

The study was conducted at the Steinberg Centre for Simulation and Interactive Learning. Each participant was present at the simulation centre for one 2.5-hour session. The simulation centre provided standardized patients (SPs), professional actors trained to act as patients. Nine SPs participated in the study and were compensated for their involvement. The use of SPs was necessary to ensure that each clinical scenario was identical, and that participants would each respond to the same scenario. The simulation centre setup included a one-way mirror arrangement and an auditory recording system. This design allowed RAs to observe and listen to the interaction between the participants and SPs.

Ten RAs were recruited and underwent training prior to the start of the study. RA training involved familiarity with the CDSS and knowledge regarding troubleshooting technological difficulties. Each RA was paired with a single participant on the three dates of the study.

Three clinical scenarios were created, each positioned along the spectrum of severity of major depressive disorder, drawing from real patient data acquired from the de-identified datasets on which the CDSS model was trained. The first patient, “Jack”, a retired Caucasian male in his 80s, suffered from mild depression, experiencing sleep disturbance, social isolation and feelings of guilt. The second patient, “Emma”, a Caucasian female in her 40s suffered a moderate depression stemming from poor work performance and emotional unavailability with her partner. The third patient, “Sara”, an African-American female in her 50s, suffered from a severe depression induced by unemployment, and demonstrated fatigue and psychomotor retardation. There were three SPs assigned to each of the three roles so that multiple simulation scenarios could take place at the same time.

Participants received an introductory presentation regarding the current state of clinical depression treatment and the basis for the development of the CDSS model. The usefulness of AI was illustrated. We explained how AI helps find complex, nonlinear patterns in data and helps us understand how many factors can influence each other and lead to remission. This model is designed to help practitioners choose the right treatments to improve results. This model was built to perform differential treatment benefit prediction. To do so, it was trained on several large datasets (STAR*D, CO-MED, REVAMP, EMBARC, and IRL-GREY). The sample was about 4700 people and was representative of a usual adult outpatients when compared to the population of Quebec. From hundreds of possible inputs, the model chose 30 key features covering 7 common treatments (medications) or treatment combinations. The baseline population remission rate is 34.85% and using this model to select treatment could improve population remission rate of 8-17%. Participants were given an overview of the tool’s user interface. They were introduced to the report page where they can access the questionnaires completed by patients. The treatment algorithm was then explained step by step. In order to get the appropriate treatment, the algorithm poses questions about the patient's condition: are they currently receiving treatment, are they suffering from psychosis, etc. A treatment list is then issued with a patient-specific remission rate. Information is available on treatments’ side effects. The physician then chooses the treatment they consider to be most appropriate. After the presentation, each participant had the opportunity to explore the tool with an RA before the simulation.

Each participant completed a demographics questionnaire, which included information about their medical training, number of years of experience, practice environments and styles, and trust in AI technology. The questionnaire also asked whether they used psychotherapy and neuromodulation technologies in the treatment of their patients with depression. Participants were informed that the “patients” they would meet had used the tool to complete questionnaires prior to their session, but had a limited understanding of how the AI model operated.

Each participant was assigned a room at the simulation centre and was provided with a laptop to access the CDSS. RAs guided their assigned participant through a ten-minute training session with the CDSS in which they directed the participants to the patient profile page, the report based on patient questionnaire responses and the treatment algorithm page. RAs responded to inquiries about the interface from the participant. All participants completed a “Station 1” questionnaire following their training with the tool to express their initial impressions and comfort with the CDSS model.

Participants engaged in three 10-minute clinical scenarios with the SPs with mild, moderate and severe depression. The order of the scenarios was random. They conducted the consultation with the SP as they would in their typical practice and integrated the CDSS model as they saw fit. The participants had access to the patient’s profile, questionnaire results, and the treatment algorithm with the integrated predictive model during each clinical scenario. The laptop with the CDSS model was angled at 45 degrees towards the participant so that the researchers could see the screen through the one-way mirror, but the participants were free to move the laptop towards themselves or towards their patient. The participants were made aware that the clinical scenarios were ten minutes in duration, and had access to a clock to keep track of the time. A study coordinator used the centre’s intercom system to remind participants when there were 3 minutes remaining to the scenario. During the scenarios, participants had the choice to use or ignore the CDSS as they saw fit. After each clinical scenario, participants completed a “Clinical Station” questionnaire regarding their use of the CDSS model during their interaction with the SP. Participants were asked about the relevance of information provided by the tool, the use of the predictive model, and their level of agreement with the treatments proposed by the CDSS.

During each clinical scenario, the assigned RA listened to and observed the interaction through the one-way mirror and the auditory recording system. RAs wrote observations regarding the participant’s behaviour with the CDSS, the physician-patient interaction, the time during which the tool was used in the clinical scenario, as well as any perceived discomfort from the participant or SP in response to the CDSS. The RA recorded any changes in the participant’s behaviour across the three clinical scenarios.

Following the three clinical scenarios, the assigned RA conducted a ten-minute interview with the participant based on their experience using the AI model in a clinical setting (see Supplementary Table 4). Participants were able to express their overall impressions of the tool, their thoughts on the accuracy of the predictive treatment model, and the potential use of the CDSS in their daily practice. Participants communicated any concerns or comments they had about the AI model, as well as ideas for improvements regarding the interface and features used.

Participants completed a final “Written Feedback” questionnaire elaborating on their use of the CDSS. They were able to give written feedback on their simulation experience and discuss future uses of the tool in their practice. Participants then completed a short quiz on the CANMAT 2016 guidelines for depression treatment in order to assess how familiar they are with these standard guidelines.

At the end of each study day, study coordinators met with the SPs to assess the impact of the CDSS on the patients’ comfort level and the ways in which the tool may influence the patient-clinician relationship from the patient’s point of view.

The data for the quantitative analysis was cleaned to remove any inconsistencies, e.g. different spellings for year of residency. The questionnaires had been created using Google Forms with pre-specified data input formats and so the number of required fixes was low. Data across questionnaires was merged by the participant’s study ID number to create a single comprehensive dataset.

**CANMAT quiz**

An independent sample t-test indicated that psychiatrists scored significantly higher (M = 6.73, SD = 2.65) on the quiz evaluating knowledge of CANMAT 2016 guidelines for depression treatment compared to PCPs (M = 4.78, SD = 1.56) (t(16.572) = -2.04, p = 0.0286). This indicates that PCPs as a group were less familiar with the standard treatment guidelines than the psychiatrists. PCPs may be more likely to welcome the assistance of the CDSS given that they are not specialized in prescribing psychiatric treatments.

**Analysis predicting physician use of CDSS suggested treatments**

Given the tool’s design to preserve physician autonomy and promote interpretability in the presentation of AI predictions, we hypothesized that physicians indicating greater trust in the CDSS would be more likely to use the remission probabilities when making treatment decisions, and that this relationship might be affected by patient’s depression severity.

A central feature of the CDSS is to predict the patient’s probability of remission with each antidepressant. To estimate the predictors of physician choice of treatments, we used multilevel modeling with mixed-effects binary logistic regression models with random intercepts at the level of the physician. The model was specified to predict whether physician treatment decisions matched one of the top two treatments outputted by the CDSS (i.e. those with the highest remision probabilities; the top two were chosen because there were often small differences in predicted percentage for these treatments, see Table 2). The main independent variables were depression severity (mild, moderate, severe) and physician trust in the CDSS, as assessed from a questionnaire administered at the end of each patient session. These variables were selected to evaluate the effect of depression severity on CDSS use and to address our hypothesis that physicians indicating greater trust in the CDSS would be more likely to use the displayed remission probabilities. Physician trust was broken down into a between- and within-subject effects. Physician-wise mean trust score (between) indicated whether physicians who were more trusting, on average, made more decisions consistent with the CDSS’s top two treatments. Mean-centered score (within) indicated whether physicians' relative trust for a given patient scenario predicted the consistency of their treatment decision with the CDSS’s top two treatments. Patient severity was divided into two dummy variables with mild depression as the reference: one representing the difference between mild and moderate depression (D1), the other difference between mild to severe depression (D2), allowing for representation of the three patient severities in the analysis. We built four models: a null model, a model for the main effects (mean trust, mean-centered trust, dummy variables for depression severity, physician ID as a random effect), a model for potential interactions between main effects, and a model for potential interactions between main effects, adding predictors of interest: physician specialty and years of experience binned (Finch et al., 2014). We used the Wald method to compute the confidence intervals for model parameters and chi-square tests to compare the fit between models (Finch et al., 2014). Analyses were performed in SPSS version 26.0.0.0 and RStudio version 1.0.136 using the (g)lmer function of the lme4 package (linear mixed-effects models). Significance was evaluated at p<0.05.

Over the three scenarios, comprising 60 individual sessions, physicians selected one of the two treatments with the highest probability of remission as predicted by the AI model 48% of the time. In order to understand what drove physician use of the treatments suggested by the AI we tested a series of four models, which are summarized in Supplementary Table 2. There was a significant interaction of severity (mild vs. severe) and mean physician-wise trust score (between-subjects factor), suggesting an additive effect between trust and severity, such that when faced with a patient presenting with more severe depression, if the physician feels more trust towards the CDSS they would show a greater tendency to use one of the treatments with the highest remission probability. There was also a significant main effect of mean-centered trust, indicating that physicians who are more trusting on average will make more treatment decisions consistent with the CDSS’s top treatment suggestions. (Supplementary Figure 1).

Since physician trust in the model was a significant term in our model both within and between-subjects, we constructed a correlation matrix as a post-hoc, exploratory analysis to investigate which variables predict physician trust in the model (Pearson correlation, 2-tailed, uncorrected p<0.05, Supplementary Figure 2). Trust in the model indicated by the feedback questionnaire at exit (representing how physicians felt about the model at the end of the study after experiencing all sessions) was significantly correlated with the degree to which physicians found the interpretability reports relevant to the patient they were treating (r=0.526, p=0.017). All other correlations with trust were non-significant.

**Discussion of analysis predicting physician use of CDSS suggested treatments**

We were interested in seeing how often physicians selected one of two treatments predicted by the AI as having the highest likely remission rate. We found that nearly half (48%) of the time, physicians chose one of the top two treatments (see supplementary materials). Given that the physicians had access to a large number of treatments (i.e. a dozen antidepressants classified as first-line treatments by the CANMAT guidelines), this appears to be a fairly high rate of agreement. Some of this may be due to the fact that the top two treatments suggested for each of the standardized patients (Supplementary Table 2) included commonly used antidepressants (i.e. citalopram, escitalopram, and sertraline). However, there were a number of commonly used antidepressants not in the top two for all three patients (i.e. mirtazapine, venlafaxine, and bupropion monotherapy). As such, there appears to be some influence - though importantly, not an overpowering one - of the AI CDSS results on clinical decision making.

We sought to understand what factors predicted physicians prescribing one of the top two treatments, and found that the predictors were physician trust in the tool and patient severity. This is a key result, because the tool’s predictions can only have their full impact when physicians use them, and so understanding these factors can help identify areas of improvement that could encourage physician use. In the simulations, physicians used the AI results more when the patient had greater depression severity and when they trusted the tool, with the two factors interacting. With respect to depression severity, it is intuitive that physicians would most seek out the support of a CDSS in their decision making in more complex cases. This is supported by the finding that 50% of physicians would use the tool for all their patients with depression, with an additional 35% noting they would reserve the tool for more severe or treatment-resistant patients (Supplementary Table 2). In addition, when a patient is less severely ill, the AI tends to predict better outcomes over a number of treatments (given the influence of depression severity as a good prognostic factor (Saragoussi et al., 2017), perhaps reducing the utility of an AI decision support in these cases. However, it is the more severely depressed and treatment resistant patients who account for a majority of resource use and poor clinical outcomes (Amos et al., 2018; Johnston et al., 2019; Mrazek et al., 2014). As such, AI decision support seems to be most acceptable to physicians where it can best be put to use.

Patient severity is not a modifiable factor over the course of a single session when a physician must make a treatment decision; however, the second predictor of physician use of the AI results, trust in the AI model, could likely be modified (Fritz and Holton, 2019). In order to understand what drove trust and therefore understand what could be improved to increase trust, we conducted an exploratory correlation analysis. The analysis revealed that the sole significant predictor of trust was the extent to which the interpretability report we provided related directly to the patient. A great deal has been written about the importance of transparency and interpretability of AI in medical tools (Benrimoh et al., 2018; Garcia-Vidal et al., 2019; He et al., 2019; Kelly et al., 2019), literature which originally motivated us to develop and include the interpretability report; this result suggests that further improvement in interpretability could directly increase physician trust in and, consequently, physician use of CDSS AI results. It is also encouraging to note that physicians, even those who are not used to working with AI, are cautious and seek to understand the tools they are working with prior to placing their trust in them and allowing them to influence their decisions. This, in turn, serves as a reminder of the importance of designing CDSS tools in collaboration with physicians, in order to take into account their concerns and to build into the tool the features required to foster trust and ensure transparency.

**Supplementary Tables and Figures**

**Supplementary Figure 1. Plot of standardized beta coefficients for variables included in the final model.** There was a significant interaction of severity (mild vs. severe) and mean physician-wise trust score (between-subjects factor) and significant main effect of mean-centered trust. In terms of years of experience, physicians with fewer years of staff experience responded with “Yes” more so than their colleagues with more years of staff experience (refer to Supplementary Table 2 for full breakdown).

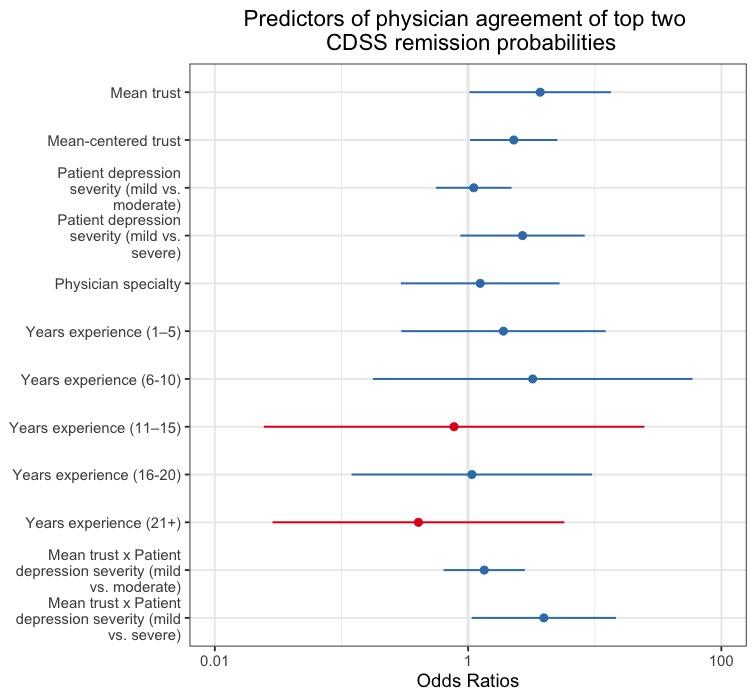

**Supplementary Table 1. Questionnaire responses (n = 20)**

| **Pre-Simulation Experience** | | |
| --- | --- | --- |
| **Question** | **Response Options** | **Frequency** |
| **Number of MDD Patients Treated Per Month**  *How often would you say you treat people with MDD (number of patients per month)?* | **0-14**  **15-29**  **30-44**  **45-59**  **60+** | **7 (35%)**  **6 (30%)**  **4 (20%)**  **1 (5%)**  **2 (10%)** |
| **Follow MDD Treatment Guidelines**  *Would you say you frequently refer back to or follow treatment guidelines when treating MDD? (1 being “not frequently” and 5 being “very frequently”)* | **1**  **2**  **3**  **4**  **5** | **4 (20%)**  **3 (15%)**  **3 (15%)**  **8 (40%)**  **2 (10%)** |
| **Psychotherapy**  *Is psychotherapy a major part of your practice?* | **Yes**  **No** | **15 (75%)**  **5 (25%)** |
| **Clinical Decision Aid Helpful**  *Do you think that a clinical decision aid for depression treatment selection would be helpful to you in your daily practice?* | **1**  **2**  **3**  **4**  **5** | **0 (0%)**  **0 (0%)**  **6 (30%)**  **8 (40%)**  **6 (30%)** |
| **Trust in AI**  *How much do you trust AI technologies (scale 1-7)?* | **1**  **2**  **3**  **4**  **5**  **6**  **7** | **0 (0%)**  **0 (0%)**  **1 (5%)**  **5 (25%)**  **9 (45%)**  **4 (20%)**  **1 (5%)** |
| **Time using clinical decision aid**  *How much time would you realistically spend using a clinical decision aid in a patient encounter or to help you make a treatment decision?* | **Less than 5 minutes**  **5 minutes**  **5 to 7 minutes**  **7 to 10 minutes**  **10-15 minutes** | **10 (50%)**  **5 (25%)**  **2 (10%)**  **2 (10%)**  **1 (5%)** |
| **Post-Simulation Experience** | | |
| **Physician use of CDSS suggested treatments**  *Did the physician prescribe one of the top two CDSS suggested treatments with highest remission probabilities? Jack: Escitalopram (81.26%), Escitalopram PLUS Buproprion XL (80.15%). Emma: Sertraline (45.91%), Citalopram (45.79%). Sara: Sertraline (30.36%), Citalopram (30.28%)* | **Jack (mild)**  **Emma (moderate)**  **Sara (severe)**  **Total** | **7 (35%)**  **9 (45%)**  **13 (65%)**  **48.33%** |
| **Psychotherapy prescribed**  *Alone or in combination with medication* | **Jack (mild)**  **Emma (moderate)**  **Sara (severe)**  **Total** | **10 (50%)**  **10 (50%)**  **10 (50%)**  **50%** |
| **Combination therapy prescribed**  *Psychotherapy in combination with medication* | **Jack (mild)**  **Emma (moderate)**  **Sara (severe)**  **Total** | **6 (30%)**  **9 (45%)**  **10 (50%)**  **41.67%** |

**Supplementary Table 2. Multilevel models for whether physician prescribed a treatment consistent with one of the CDSS top two treatments, ranked by remission percent probability**

| Parameter | Model 1 |  | Model 2 |  | Model 3 |  | Model 4 |  |
| --- | --- | --- | --- | --- | --- | --- | --- | --- |
|  | Standardized beta (SD) | OR (CI) | Standardized beta (SD) | OR (CI) | Standardized beta (SD) | OR (CI) | Standardized beta (SD) | OR (CI) |
| Intercept | -0.067 (0.258) | 0.935 (-0.573 – 0.440) | -4.539 (1.580) | 0.011 (0.000 – 0.191) | -1.415 (2.263) | 0.243 (0.002 – 19.124 | -1.422 (3.421) | 0.241 (0.001 – 56.644) |
| Mean trust |  |  | **1.161 (0.424)**** | **3.195 (1.463 – 7.935)**** | 0.271 (0.641) | 1.312 (0.370 – 4.964) | 0.180 (0.911) | 1.197 (0.213 – 6.534) |
| Mean- centered trust |  |  | 0.901 (0.513) | 2.461 (0.958 – 7.495) | **1.266 (0.598)*** | **3.548 (1.219 – 13.324)*** | **1.302 (0.636)*** | **3.676 (1.221 – 14.728)*** |
| Patient depression severity (mild vs. moderate) |  |  | 0.337 (0.728) | 1.401 (0.335 – 6.043) | -2.282 (3.361) | 0.102 (0.000 – 70.260) | -2.444 (3.533) | 0.087 (0.000 – 29.470) |
| Patient depression severity (mild vs. severe) |  |  | 1.290 (0.754) | 3.633 (0.862 – 17.195) | -10.221 (5.421) | 0.000 (0.000 – 0.342) | -10.405 (5.477)* | 0.000 (0.000 – 0.140)* |
| Mean trust x Patient depression severity (mild vs. moderate) |  |  |  |  | 0.726 (0.954) | 2.067 (0.327 – 14.803) | 0.794 (1.020) | 2.212 (0.350 – 11.163) |
| Mean trust x Patient depression severity (mild vs. severe) |  |  |  |  | **3.683 (1.819)*** | **39.782 (2.179 – 5322.178)*** | **3.728 (1.812)*** | **41.576 (2.538 – 3180.289)*** |
| Physician specialty |  |  |  |  |  |  | 0.220 (0.736) | 1.246 (0.298 – 5.550) |
| Years experience (1–5) |  |  |  |  |  |  | 0.642 (0.949) | 1.901 (0.309 – 13.108) |
| Years experience (6–10) |  |  |  |  |  |  | 1.174 (1.482) | 3.236 (0.207 – 100.888) |
| Years experience (11–15) |  |  |  |  |  |  | -0.255 (1.765) | 0.775 (0.017 – 23.845) |
| Years experience (16–20) |  |  |  |  |  |  | 0.069 (1.116) | 1.071 (0.121 – 10.975) |
| Years experience (21+) |  |  |  |  |  |  | -0.903 (1.354) | 0.405 (0.021 – 5.224) |

Physician specialty is psychiatrist compared to the reference (PCP). The reference category for years experience is 0 (i.e., residents). Beta estimates (standard error) are shown for fixed effects and variance (standard deviation) is shown for random effect.

* p<0.05; ** p<0.01. OR: Odds Ratio; SD.: Standard Deviation; CI: Confidence Intervals

**Supplementary Table 3. Qualitative reference table for themes and sub-headings**

| Theme | Sub-Headings |
| --- | --- |
| 1. Interpretability of the Tool | 1.1 Report is helpful/useful  1.2 Report is not helpful/useful |
| 2. Impact on Treatment Decision and Clinical Practice | 2.1 Impact on tx decision: positive, negative, neutral  2.2 Probabilities of remission  2.3 Patient choice for use with AI  2.4 Openness to prescribing drugs  2.5 Added value of the tool vs. no added value  2.6 Other uses of tool  2.7 When doc would use tool and why? |
| 3. Trust, Understanding, Feelings about AI | 3.1 Request for evidence behind AI  3.2 Belief/disbelief/uncertainty  3.3 Understanding of AI  3.4 How physician explains AI / effort to explain  3.5 Concerns about bias or accuracy of data  3.6 Terminology used with patient  3.7 Physician curiosity about AI |
| 4. Impact on Physician-Patient Interaction | 4.1 Barrier to flow/facilitator of flow  4.2 Self-reported comfort with tool  4.3 Perceived comfort of physician  4.4 Impact of time spent using tool on comfort / training with the tool / use before meeting patient  4.5 Physician invited/engaged them with tool / shared tool with patient  4.6 Degree patient felt heard, trust, connection with doc  4.7 Body language and eye contact, amount of time spent using tool, silences while reading tool, how info was communicated to patient, and balance of using tool vs. interviewing |

**Supplementary Figure 2. Sunburst diagram representing relative proportions of qualitative data for themes and sub-headings**

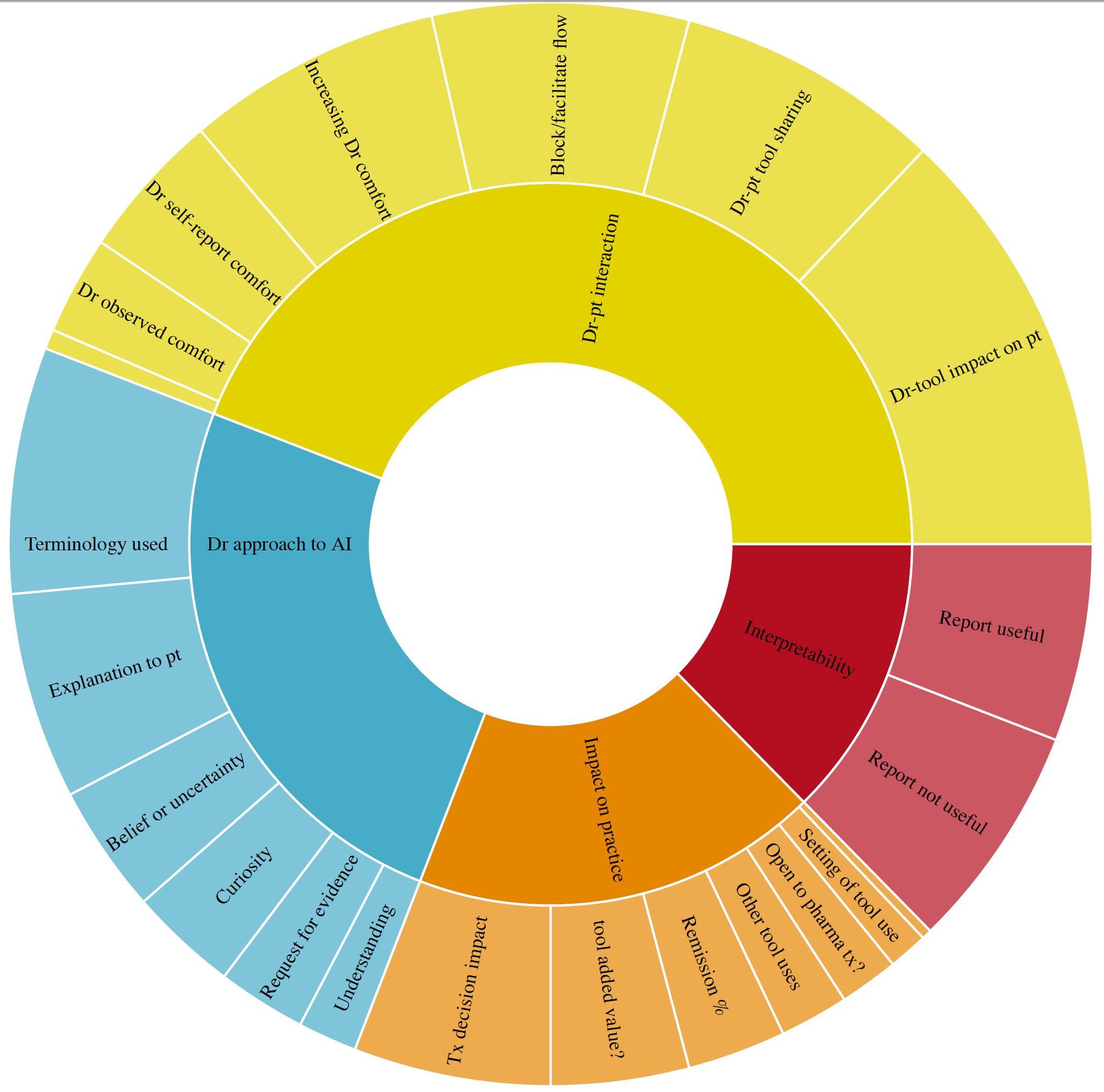

**Supplementary Table 4. Structure Interview Script** (free text entry by RAs for all questions)

**Questions posed to physicians following all three scenarios**

What was your overall impression of the application?

How did you feel the application affected the clinician-patient relationship?

How did you feel the predictive model handled the clinical situations presented?

What features of the tool did you find useful? Less useful?

Do you think you would use the application in your daily practice? If not, why not?

What could be done to make the predictive model better?

How did you find the information listed under "more info" in the predictive model (i.e. the interpretability report)?

**Supplementary Figure 3. Patient populations with whom physicians would use the CDSS**

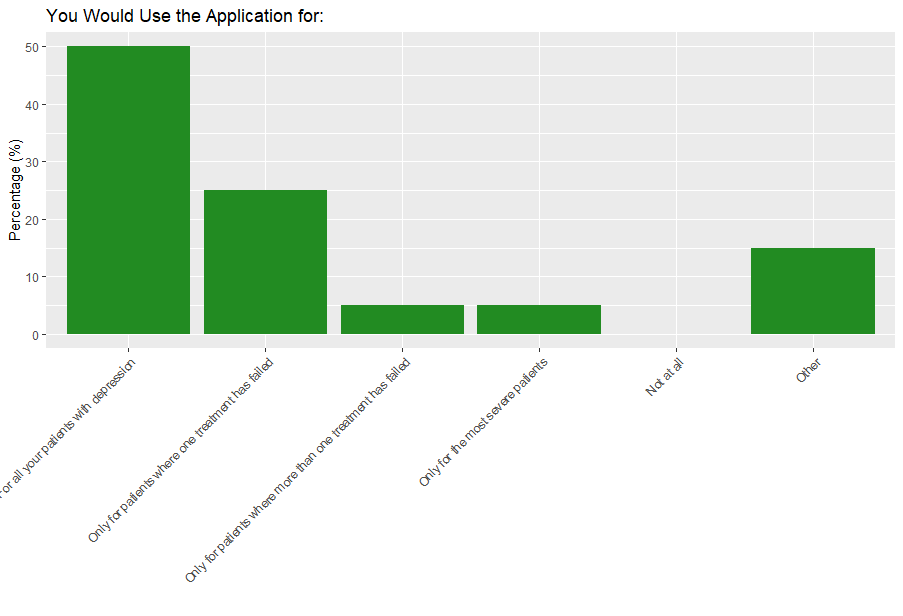

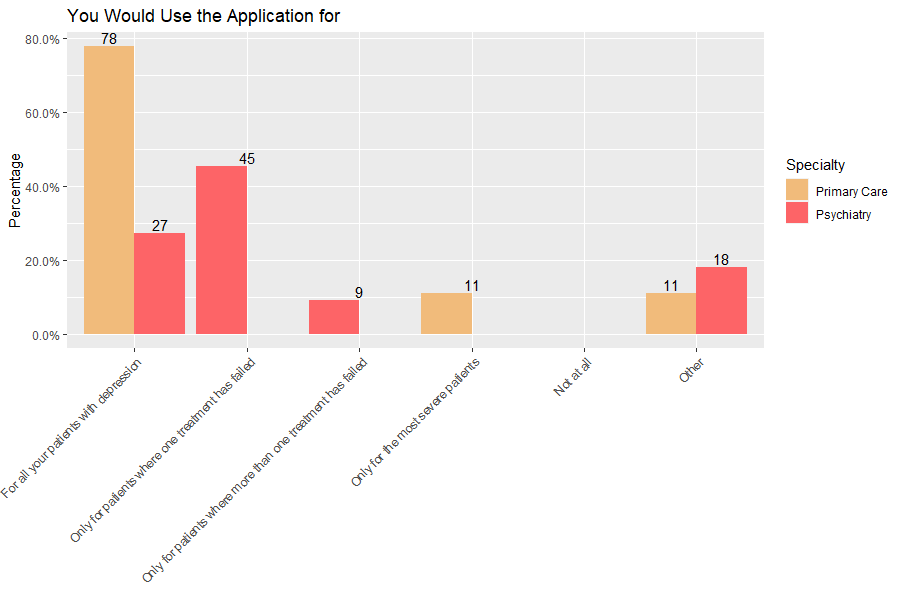
